# Infrastructure risk factors for leptospirosis transmission in urban informal settlements

**DOI:** 10.64898/2026.08.13.26360375

**Authors:** Ana Maria Nascimento Silva, Juliet Oliveira Santana, George Gonçalves Machado, Fábio Neves Souza, Daiana Santos de Oliveira, Fabiana Almerinda G. Palma, Priscilla Elizabeth Ferreira dos Santos, Paulo Ricardo Dias Pimentel, Cleber Cremonese, Federico Costa, Rodolfo B. Nobrega, Guy Howard

## Abstract

Leptospirosis is a globally important environmentally transmitted disease, with approximately one million cases and 60,000 deaths reported annually. In low-income urban communities, inadequate sanitation, drainage and waste management may increase human exposure to contaminated environments. This study investigated the influence of environmental engineering risk factors on *Leptospira* exposure in four disadvantaged urban communities (favelas) in Salvador, Brazil. A high-precision georeferenced field survey was developed to identify, map and characterise sanitation, stormwater drainage and solid waste infrastructure. Cross-sectional spatial analyses of baseline data were used to assess associations between environmental risk factors and the residential locations of individuals with anti-*Leptospira* antibodies. Seropositive individuals tended to reside closer to environmental risk factors and at lower relative elevations. Density analyses indicated that seropositive individuals tended to reside closer to inadequate or partially adequate sewerage components than to sewage-contaminated streams or open sewage points. Inadequate streets showed also showed high density peaks, suggesting that exposure may occur through frequent contact with contaminated runoff and standing water. In contrast, open waste dumping sites and vacant lots showed weaker and more diffuse spatial patterns. The findings highlight the importance of infrastructure quality shaping leptospirosis risk within urban informal settlements. The proposed methodology provides a practical approach for high-resolution characterisation of environmental exposure pathways and may support targeted engineering interventions and epidemiological investigations of leptospirosis and other environmentally transmitted diseases.

**AUTHOR SUMMARY:** Leptospirosis is transmitted through contact with water or soil contaminated by pathogenic *Leptospira*. In densely populated urban communities, inadequate sanitation, drainage, streets and waste management may create multiple exposure pathways. However, methods for assessing these conditions in sufficient detail remain limited. We developed a systematic field methodology using high-precision georeferencing to identify, map and assess environmental engineering risk factors in four disadvantaged urban communities in Salvador, Brazil. We examined these conditions in relation to the homes of residents with evidence of previous exposure to *Leptospira* bacteria. Previously exposed residents tended to live closer to inadequate or partially adequate sewerage components and inadequate streets, and at lower elevations relative to their surroundings. Waste-disposal sites and vacant lots showed less distinct patterns. Our findings show that detailed, high-precision mapping of infrastructure conditions can identify potential environmental exposure pathways that broader assessments of sanitation may overlook. This practical and reproducible methodology can support epidemiological studies of leptospirosis and other diseases transmitted through contaminated environments and help identify locations where infrastructure improvements may reduce human exposure.

## 1. INTRODUCTION

The increasing global burden of diseases associated with inadequate urban infrastructure and poor environmental sanitation is disproportionately high in low- and middle-income countries (LMICs), where leptospirosis remains an important public health concern [1,2,3]. Approximately one million cases and 60,000 deaths occur annually worldwide [4]. Human infection occurs through contact with water or soil contaminated by pathogenic *Leptospira* [5]. Low-income, inadequate sanitation, drainage, and solid waste management create favourable conditions for rat populations and environmental contamination, consequently, vulnerable urban communities are disproportionately affected by leptospirosis and other diseases [6,7].

This is particularly evident in Brazil, where historical, political, and socio-economic factors have driven unequal urban development. For example, the city of Salvador has 42.7% of its 2.4 million inhabitants living in urban low-income communities named *favelas* [8] with insufficient water, sanitation, drainage, and solid waste management [9]. In *favelas* of Salvador, Brazil, the *Leptospira* infection rate reaches 37.8 per 1,000 person-years, with an annual incidence for severe leptospirosis at 19.8 cases per 100,000 inhabitants, and where every additional US$1 rise in the daily per capita household income reduced the odds of primary infection by half [10].

Although inadequate sanitation and flooding are recognised risk factors for leptospirosis [11, 12], the specific role of sewerage and stormwater drainage components to environmental exposure remains poorly understood [13,14]. Potential risk factors beyond open sewers, such as broken or obstructed sewerage and drainage components and leaks may also contribute to exposure to contaminated water and, consequently, influence leptospirosis transmission.

The multifactorial nature of leptospirosis transmission presents important challenges for environmental exposure assessment. Infrastructure must be characterised with sufficient spatial precision to support epidemiological analyses, regarding that risk factors operate at multiple spatial scales [15]. This is critical in dense urban environments, where potential sources of contamination and human populations are closely situated. Studies of Brazilian *favelas* indicate significant spatial variability of environmental and social characteristics within these areas, also underscoring the need for high spatial precision investigations [10,16,17,18].

Previous studies have proposed methods for characterising deficiencies in urban infrastructure. However, existing approaches often lack detailed operational definitions, integration with GIS tools, or sufficient methodological information to enable replication [19,20,21] Also, existing studies typically rely on household questionnaires, remote sensing, municipal records that frequently are not available for low-income urban communities. Similarly, epidemiological studies frequently rely on environmental proxies such as polluted streams or open sewers [17], which may not adequately capture the diversity of infrastructure failures associated with environmental exposure [22].

This study set out to investigate how potential environmental engineering risk factors – specifically sanitation, storm drainage and solid waste – influence exposure to *Leptospira* in favelas with high disease burdens. The study specifically aims to (i) develop a practical, high-precision georeferenced survey for identifying and mapping urban environmental infrastructure; (ii) describe and characterise fine-scale environmental risk factors; and (iii) examine their relationship with the seroprevalence of anti-*Leptospira* antibodies. The proposed methodology may support epidemiological investigations in low-income urban communities and the designing of targeted engineering and public health interventions.

## 2. METHODS

### 2.1. Study area

This study was conducted in four urban areas in Salvador, Northeastern Brazil. Salvador’s climate is tropical humid to subhumid, with distinct wet and dry seasons. Between 2007 and 2022, 2,359 leptospirosis cases were reported in Salvador [23]. This study comprised a cross-sectional spatial analysis of baseline data collected as part of a longitudinal quasi-experimental epidemiological study [14]. The study areas were low-income urban communities selected according to criteria described previously [14]. These included high-risk areas averaging 30,000 m², where households are located within 40 m of streams contaminated by sewage and solid waste, similar topographic and hydrological characteristics, similar socio-economic status, similar population density and *Leptospira* seroprevalence.

The study areas (Fig 1) are located in the neighbourhoods of Nova Sussuarana (study area 1), Arenoso (study area 2), Jardim Santo Inácio (study area 3), and Calabetão (study area 4). These areas are in valleys with streams heavily polluted by sewage and solid waste. They are characterised by high population density, poor sanitation conditions, with aging storm drainage infrastructure that does not cover all the streets in the study areas. Some households are located on steep slopes or directly adjacent to streams, often in flood-prone zones.

**Fig 1.**
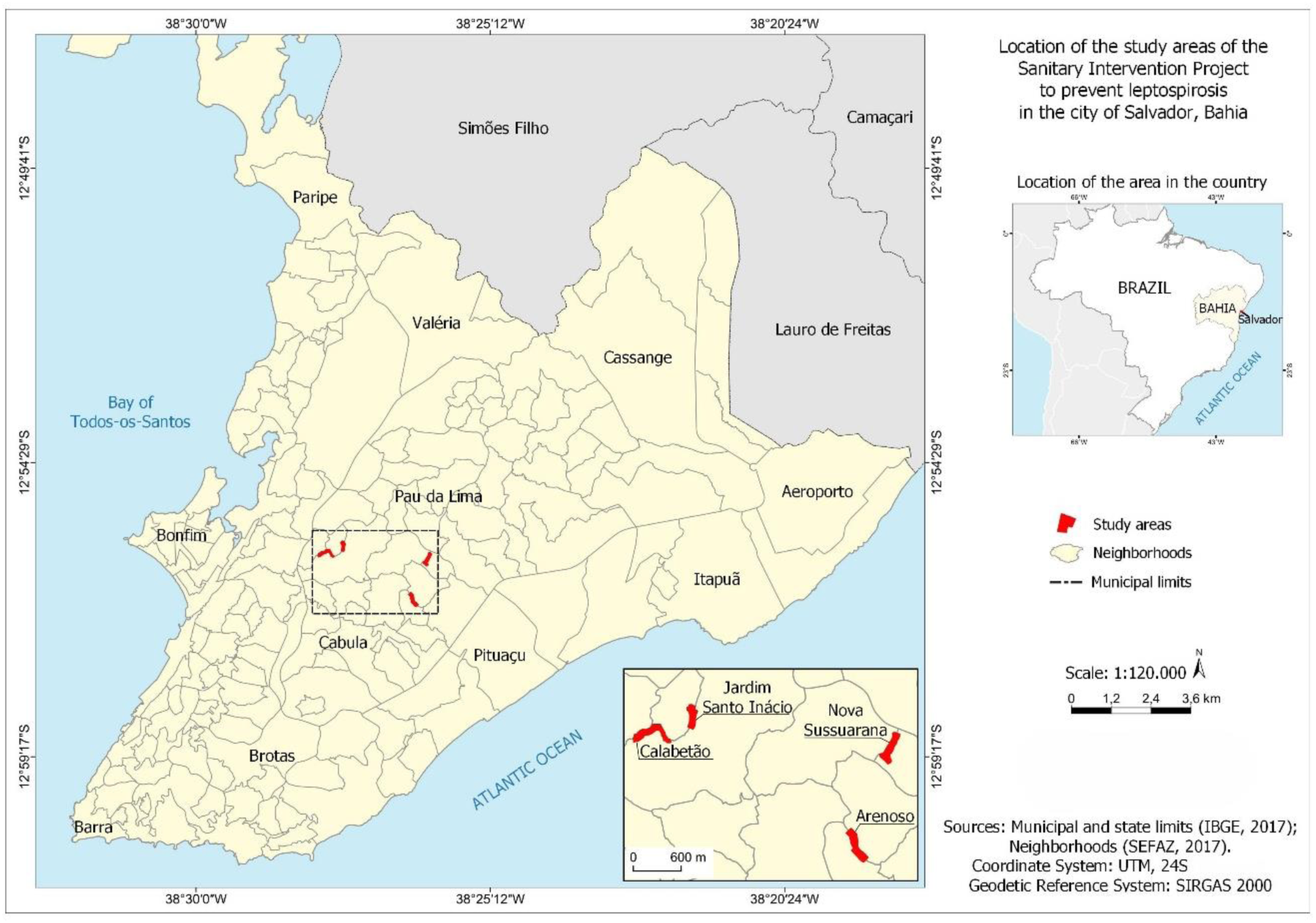
Study areas.

### 2.2. Development of data collection survey and study variables

The data collection survey was developed by an experienced multidisciplinary research team comprising environmental engineers, epidemiologists, geographers, ecologists, and public health specialists. The research team has been working in Salvador’s low-income urban communities for years. The development process began with the elaboration of a structured questionnaire, followed by a series of meetings to discuss its content, format, theoretical and operational categories, until consensus was achieved. The instrument was reviewed during meetings with an external specialist in urban infrastructure and environmental sanitation, who had previously developed a framework for local urban environmental quality assessment [19]. Feedback from these meetings informed the refinement of the field survey.

Preliminary field visits were undertaken to identify key infrastructure issues such as the partial coverage of households by sewerage systems, ageing drainage systems, and inadequate solid waste management. The final survey was systematised into seven standardised forms, containing 40 questions, which was piloted, and implemented in REDcap^®^ [24]. The instrument allowed the identification and characterisation of the variables detailed in Table 1.

**Table 1.**
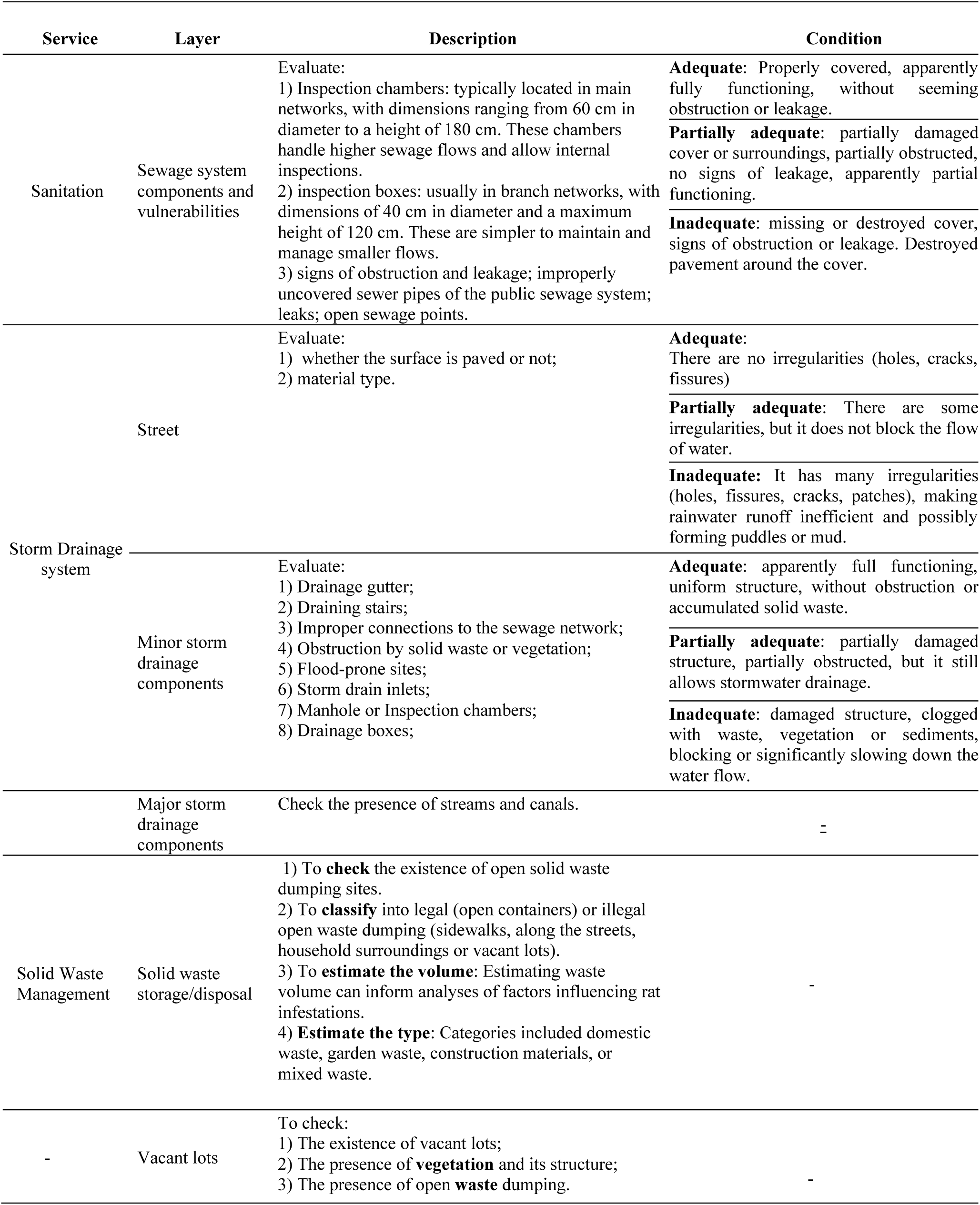
Study variables.

The field procedures were systematised in a data collection protocol by the multidisciplinary research team to ensure consistency during fieldwork [25]. The protocol was based on the discussions, field observations, literature review, and standards adopted by the local public sanitation company. It described the definitions of each variable, assessment criteria, provide photographic examples, and georeferencing procedures. The protocol was refined through pilot testing to improve clarity of assessment criteria and procedures.

The data collection survey was designed to identify and characterise points of exposure related to sanitation, stormwater drainage, and solid waste management, using high precision georeferencing (≤ 50 cm). The study areas feature relatively short distances between households and potential contamination sources. Accurate geolocation was critical to capturing these spatial nuances. To reduce the risks of misplacement errors that can arise from the use of a Global Positioning System (GPS), a Global Navigation Satellite System (GNSS) Spectra Geospatial SP60 was used as this has access to other systems beyond GPS, such as GLONASS and Beidou [26].

Infrastructure condition classification as adequate, partially adequate, and inadequate was based on and adapted from the urban environmental quality assessment framework proposed by [19]. Infrastructure considered to be in an adequate condition represents the preferred condition where infrastructure components are functioning properly. Inadequate indicates components in poor condition that potentially pose a higher risk for human contact with contaminated water or soil and may act as sources of contamination. Partially adequate refers to components that are between the two extremes – those that have not yet failed but show signs of potential failure in the short term. Without timely intervention, components in this category may deteriorate and become inadequate. The variables for solid waste and vacant lots did not include assessment of condition but assessment focused on identifying open waste dumping sites and vacant lots, as their presence indicates inadequacy in waste management. Similarly, the stream in each study area was identified but not assessed for condition, as its presence itself was considered a risk factor [16,17].

Drainage components with considerable longitudinal dimensions (e.g. gutters and stairs) were measured in 10–30 m segments, and the same was done for streets, ensuring comprehensive spatial coverage while maintaining data collection efficiency. The condition of all components was evaluated based on their capacity to drain stormwater and reduce human contact with contaminated water. The segments were defined based on three criteria: length, material (asphalt, exposed soil, concrete and cement), and condition (S1 Table). The segment length was set at a maximum of 30 m to capture sufficient detail and 10 m set as the minimum and lengths shorter than this would have substantially increased the workload without providing significant additional insights.

### 2.3. Data collection

Data collection was conducted between June 2022 and January 2023 across the four study areas. Using printed maps, the process began by systematically walking through each street to characterise components by visual inspection. Observations were classified according to the procedures described in the protocol and recorded using the REDCap® forms. Although there is no exact number of deficiencies (e.g. fissures and cracks) that differentiates partially adequate to inadequate status, we sought to minimise the subjectivity of the assessment. The field assessments followed the standardized data collection protocol in all study areas.

Field assessments were conducted by a team comprising at least an environmental engineer (infrastructure assessment), a geographer (georeferencing), a field assistant (photographic documentation) and a community engagement agent (local guidance). To ensure consistency, the initial field survey was conducted jointly by two environmental engineers, allowing them to align the application of the assessment criteria. Likewise, two geographers performed the initial survey together to standardize the georeferencing procedures.

The survey was applied to formal and informal streets of the communities, encompassing all alleys, stairs, and paths not represented on official maps. This ensured we could map all the infrastructure present, because informal paths and streets in these settings often lack or have insufficient basic services and may serve as hidden sources of contamination. Focusing solely on formal streets could overlook key data relevant to leptospirosis transmission. Additionally, informal streets are typically absent from detailed satellite imagery and are not mapped by platforms like Google Street View. The comprehensive mapping we adopted offers a novel contribution to leptospirosis research and broader environmental studies in urban settings. Each characterised component had its coordinates collected using a Spectra Geospatial SP60 GNSS receiver. Photos of each georeferenced component were taken.

### 2.4. Data quality control

Data quality control was conducted both during and after fieldwork by the engineer and the geographer. This process involved verifying that all characterised components matched the georeferenced data, using the component ID to cross-reference data from different sources (REDCap®, GNSS, and photo files). The first step was to verify that the number of components matched across the datasets, particularly between REDCap® and GNSS. While it was preferable to have a corresponding photo for each component, missing photos were not critical to data analyses. The second step in data quality control involved verifying the status of the REDCap® forms. Each form included a checkbox indicating whether it was marked as incomplete, complete, or unverified. Incomplete or unverified forms were reviewed, and associated photographs and georeferenced locations were used to fill in missing fields as far as possible. If photos were insufficient to complete the information accurately, a further field visit was undertaken to collect the missing data. The process of data quality control is further detailed in Fig 2.

**Fig 2.**
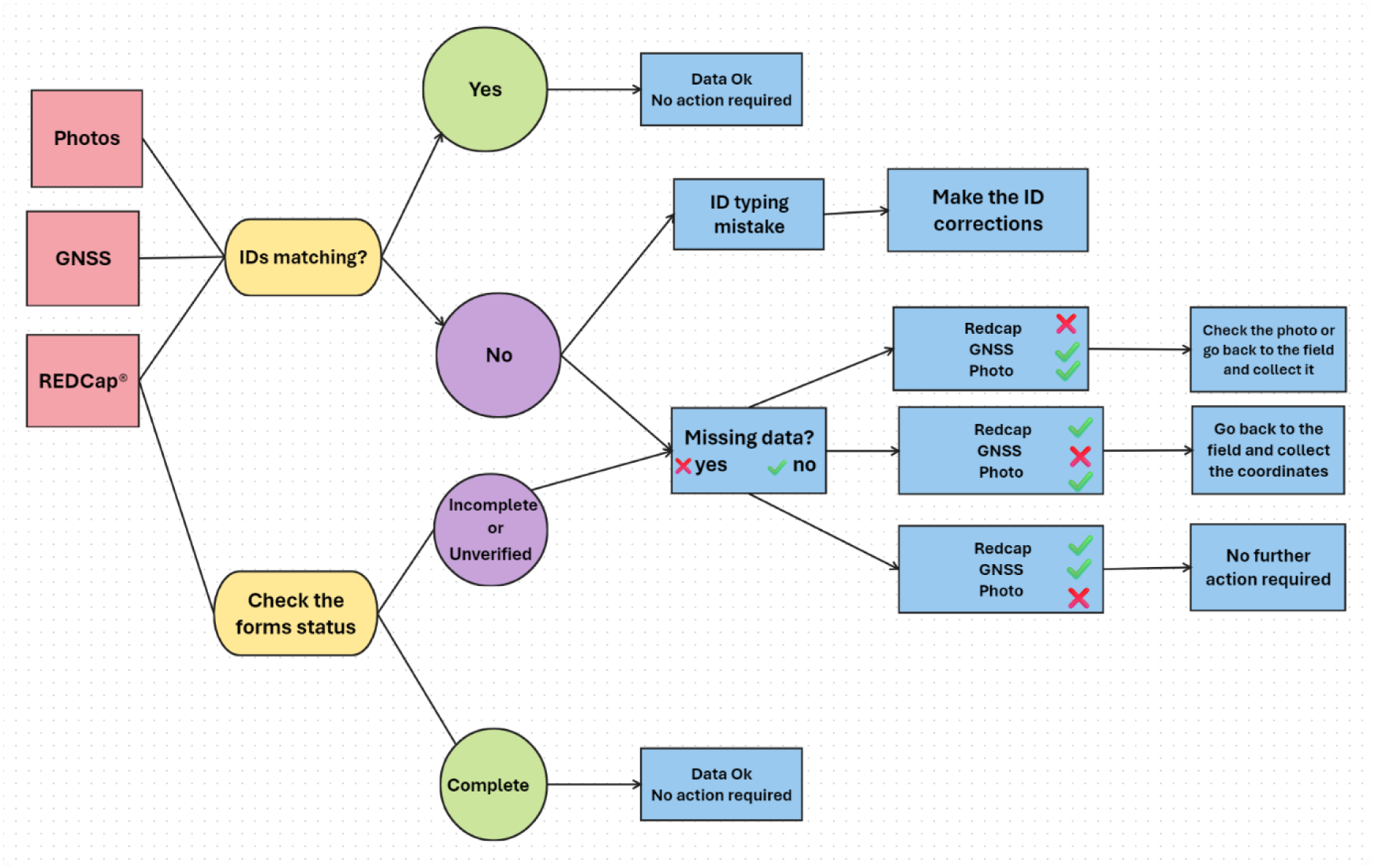
Data quality control process.

### 2.5. Data analyses

#### 2.5.1. Selection of Potential Risk Factors

Seven risk factors were selected based on variables (Table 1) indicating the potential for human exposure to contaminated water and soil. Regarding sanitation, the selected risk factors were the open sewage points and partially adequate or inadequate sewerage components. For drainage, partially adequate and inadequate components, streams, and inadequate streets. Open waste dumping sites and vacant lots were also deemed as potential risk factors.

#### 2.5.2. Spatialising the Data

Once the environmental engineering risk factors were selected, the next step involved spatialising and cleaning the data in ArcGIS 10.7.1 [27] to prevent overlapping. This step ensures that each risk factor is counted only once. For instance, open sewage points were separated from streams and inadequate sewerage components. By definition, open sewage points refer to domestic sewage discharged into the environment, resulting in pools of contaminated water. These were not considered if they originated from inadequate sewerage or drainage systems or coincided with the stream. In cases where an inadequate sewerage component pours contaminated water into the environment, it was classified as an inadequate sewerage component alone, not as an open sewage point, to avoid double counting.

The shapefiles containing the streams that pass through Nova Sussuarana and Arenoso were obtained from the Salvador City Hall [28] and for Jardim Santo Inacio and Calabetao were provided by The Urban Development Company of Bahia [29]. Additionally, coordinates of some points along the streams were collected using high precision georeferencing to ensure streams lines and points were matching.

#### 2.5.3. Case-study: Seroprevalence of anti-*Leptospira* antibodies and its spatial distribution

Serological data used in this study were obtained from an ongoing population-based serological survey conducted in the study communities in Salvador, Brazil [14]. Between 16^th^ September 2021 and 16^th^ December 2022, a total of 1,075 participants were recruited. The mean age was 32 years, with 58.1% female and 41.9% male. Participants were distributed across the four study areas: 297 in Nova Sussuarana, 237 in Arenoso, 277 in Jardim Santo Inácio and 264 in Calabetão. No separate sample-size calculation was undertaken for the present cross-sectional analysis, all participants with available serological results and valid geocoded residential data were included. Following written informed consent, trained healthcare professionals collected venous blood samples during household visits. Samples were transported under refrigerated conditions to the Laboratory of Gonçalo Moniz Institute, Oswaldo Cruz Foundation (Fiocruz Bahia), where serum was separated by centrifugation, aliquoted, and stored at −20°C until serological analysis.

Serum samples were tested for anti-*Leptospira* antibodies using the microscopic agglutination test (MAT), the reference serological assay for leptospirosis. MAT was performed using a panel of live *Leptospira* serovars representative of the major pathogenic serogroups circulating in Brazil, following previously described protocols [30]. Samples were initially screened at a 1:50 dilution, and those exhibiting at least 50% agglutination were considered reactive. Reactive samples were subsequently titrated in serial two-fold dilutions to determine the endpoint antibody titer. Participants with MAT titers ≥1:50 were classified as seropositive. For seropositive samples, the presumptive infecting serogroup was assigned according to the highest MAT titer, while samples with identical highest titers for more than one serogroup were classified as mixed serogroup reactions, as previously described [30].

To quantify the individual-level of environmental exposure, the Generate Near Table tool in ArcGIS 10.7.1 [27] was used to calculate the shortest distance between each household and each mapped potential sanitary and environmental risk factor.

Density plots explored the spatial distribution of individuals with anti-*Leptospira* antibodies concerning their nearest distances to environmental risk factors and the relative elevation of households to the lowest point of each study area. These plots were generated for each study area and its sub-areas. Dividing study areas into sub-areas aimed to identify potential variability influencing exposure to leptospirosis risk factors. Sub-areas were delineated based on hydrological features, given the potential relationship between *Leptospira* infections and hydrological patterns [31].

The division was made based on sub-catchments-like areas within each catchment, defined by terrain elevation, land use, and street networks, including informal streets, resulting in upper, intermediate, and lower sub-catchments (Table 2, S1 Fig). Sub-areas for each study area were adjusted to have similar sizes (within a 20% difference) to facilitate comparative analysis. Sub-areas size varies from 1.02 to 1.92 ha across all study areas.

**Table 2.** Sub-areas definition across the study areas.

| Subareas | Study Areas |  |  |  |
| --- | --- | --- | --- | --- |
|  | Nova Sussuarana | Arenoso | Jardim Santo Inacio | Calabetao |
| Upper | NS HIGH | ARE HIGH | JSI HIGH | CAL HIGH |
| Intermediate | NS MID | ARE MID | JSI MID | CAL MID |
| Lower | NS LOW | ARE LOW | JSI LOW | CAL LOW |

After defining sub-areas, distances to environmental risk factors were organised accordingly. Kernel density estimation (KDE) was used to generate density plots in R version 4.4.1 [32] using the geom_density function from ggplot2 package [33]. By default, R employs the Gaussian kernel, with the bandwidth as the standard deviation of the smoothing kernel without weights. This results in density plots where the area under the curve equals 1, and the highest density values represent the most probable values for each variable.

### 2.6. Ethics

Ethical approval for this study was obtained from the ethics committee at the Collective Health Institute, Federal University of Bahia (CEP/ISC/UFBA) under number CAEE 32361820.7.0000.5030, and the National Research Ethics Committee (CONEP) linked to the Brazilian Ministry of Health under approval number 4.235.251. All participants involved in the study provided written informed consent before data collection.

## 3. RESULTS

### 3.1. Spatial distribution of seroprevalence and potential risk factors

Overall, between 35% and 64% of the streets are in inadequate condition across the study areas Fig 3 frequently worsening from uphill to downhill (Fig 4a,b,d). Inadequate streets consistently showed among the highest density peaks and shortest distances to individuals with anti-Leptospira antibodies, with density peaks generally within 5 and 10 m (Table 3). Across sub-areas, density patterns for inadequate streets varied between study areas, with no consistent upstream–downstream trend (Fig 5).

**Fig 3.**
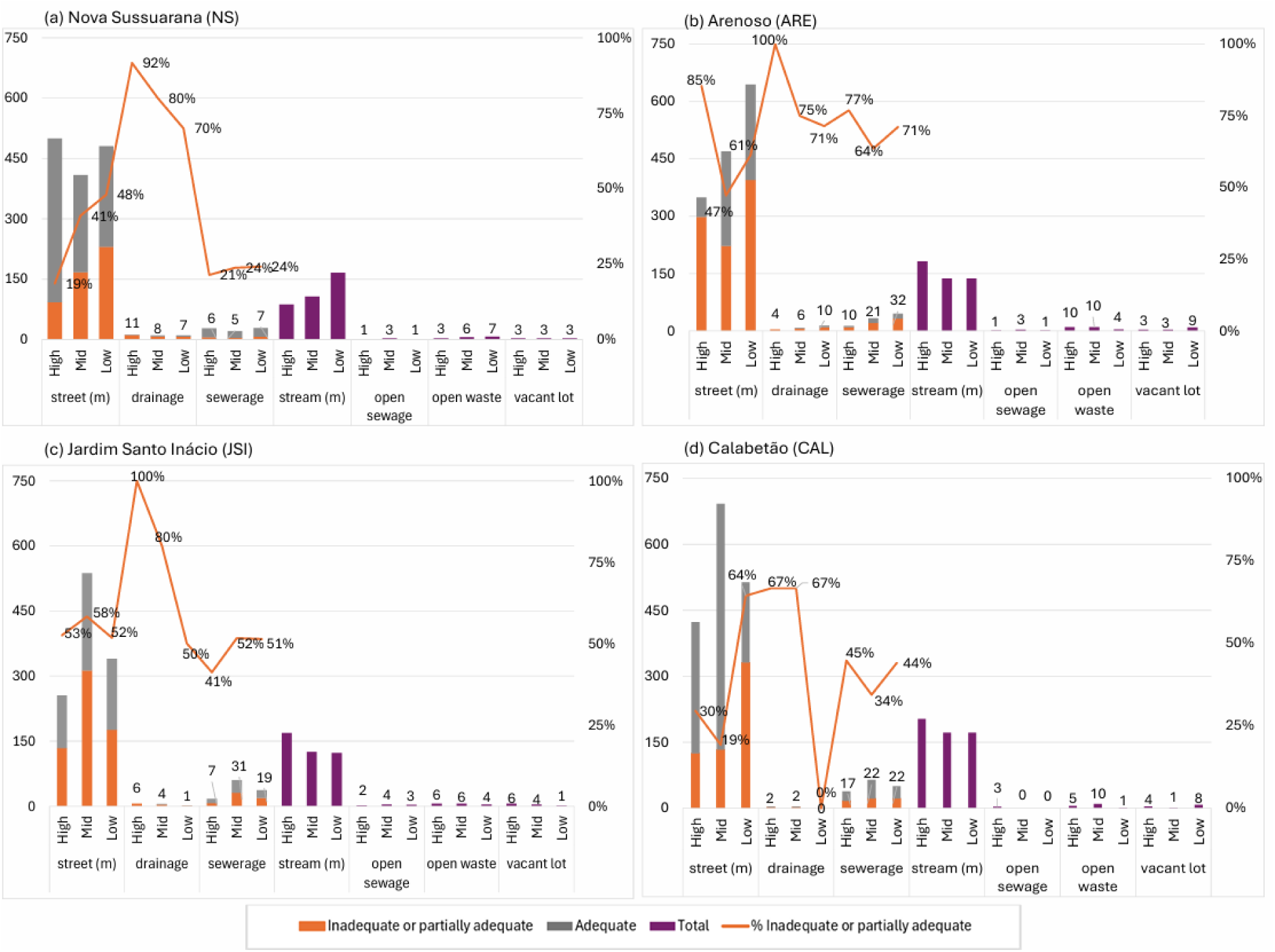
Distribution and condition of mapped environmental infrastructure across study areas and sub-areas.

**Fig 4.**
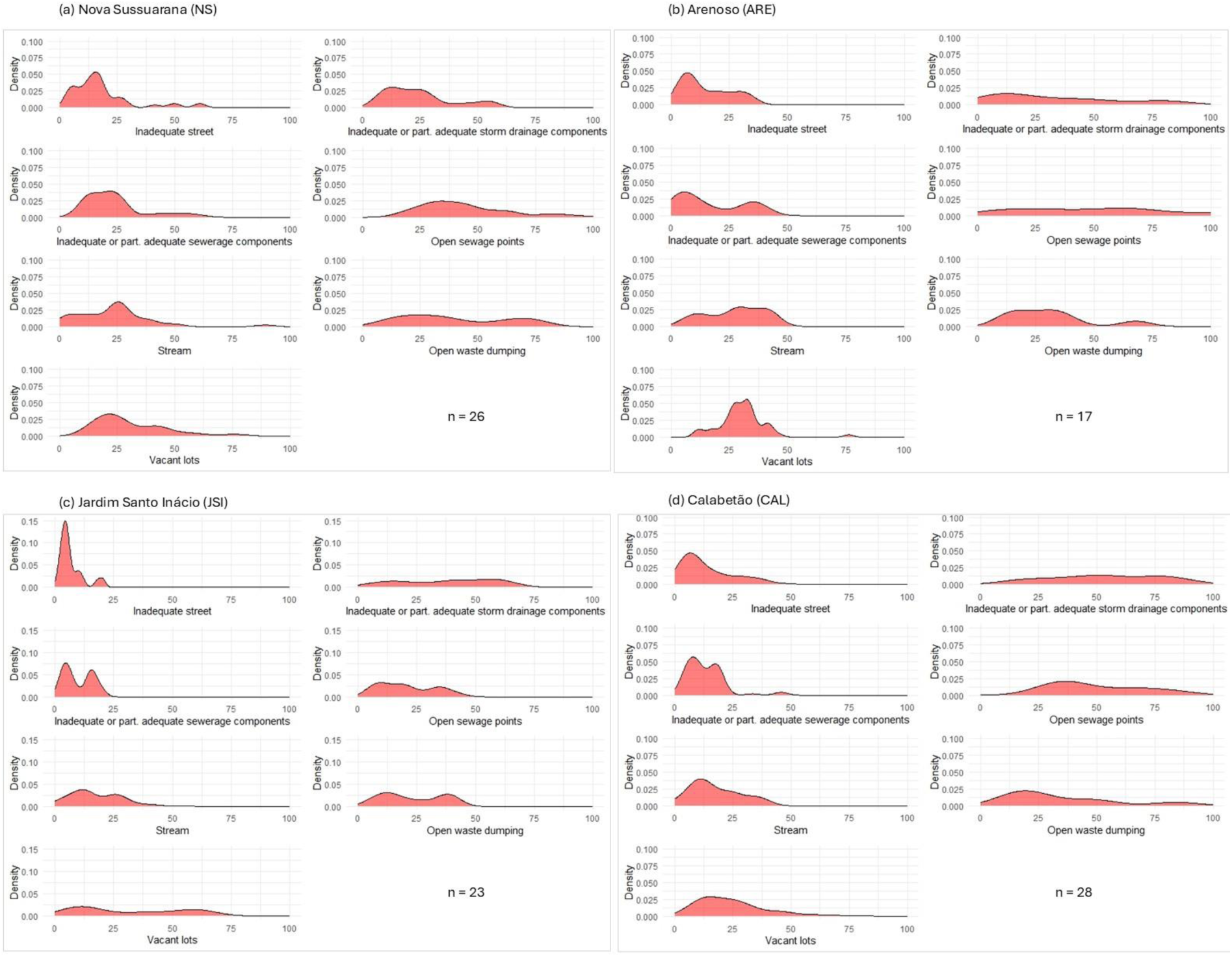
Probability density of the distance (m) between seropositive individuals and potential environmental risk factors in each study area. Panels (a-d) correspond to the four study areas. *n* indicates the number of seropositive individuals in each study area. Among 1,075 participants with available serological results 94 (8.7%) were seropositive for anti-*Leptospira* antibodies.

**Fig 5.**
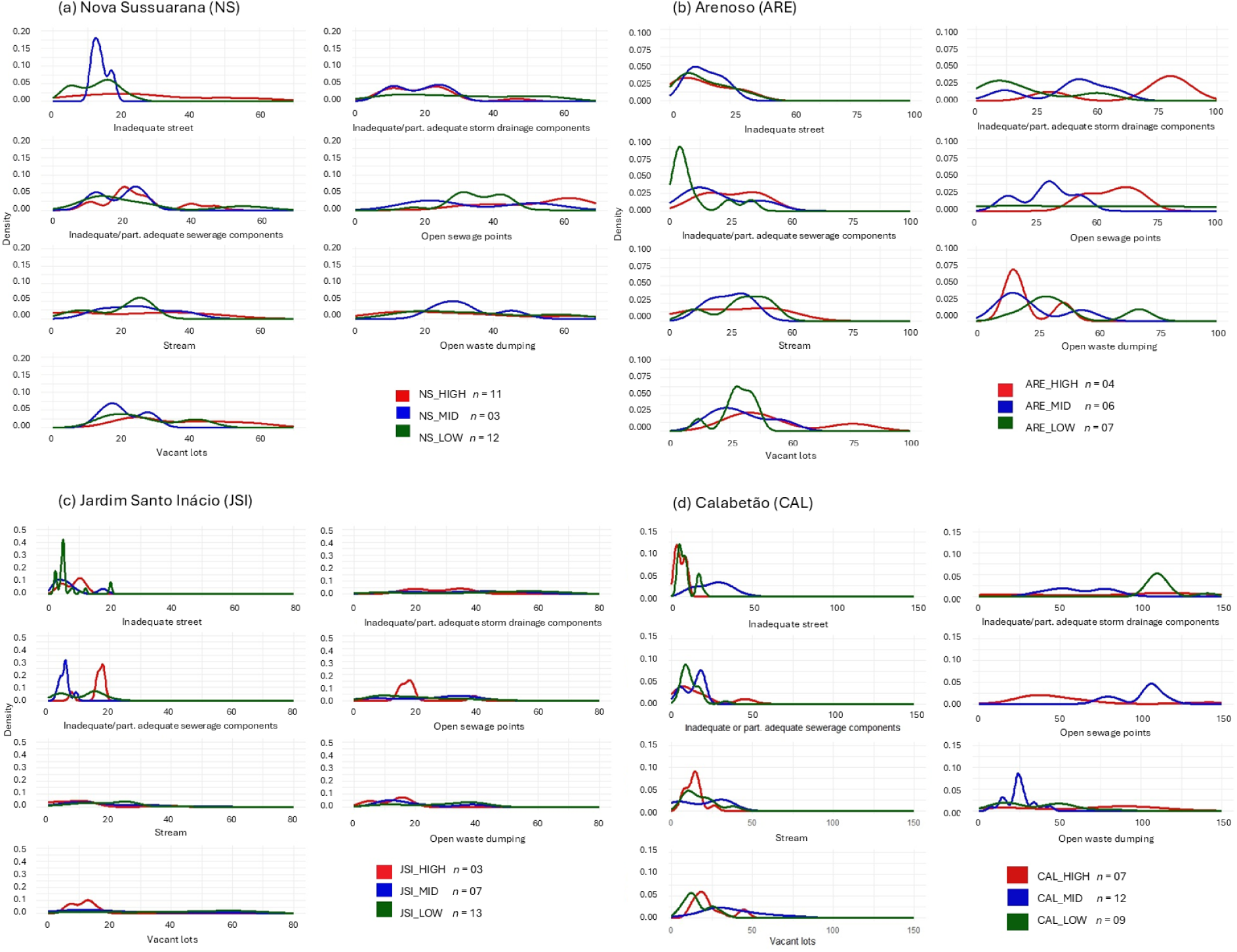
Probability density of the distance (m) between seropositive individuals and potential environmental risk factors by sub-area within each study area. Panels (a-d) correspond to each study area. *n* = number of seropositive individuals in each subarea.

**Table 3.** Peak densities and distances by potential risk factors.

|  | Inadequate Street |  | Inadequate or part. adeq. Drainage |  | Inadequate or part. adeq. Sewerage |  | Stream |  | Open sewage points |  | Open waste dumping sites |  | Vacant lots |  |
| --- | --- | --- | --- | --- | --- | --- | --- | --- | --- | --- | --- | --- | --- | --- |
|  | DP | PD (m) | DP | PD (m) | DP | PD (m) | DP | PD (m) | DP | PD (m) | DP | PD (m) | DP | PD (m) |
| **NS** | **0.050** | **15** | 0.030 | 13-25 | 0.040 | 13-25 | 0.040 | 25 | NP | NP | NP | NP | 0.035 | 20 |
| **NS_HIGH** | 0.025 | 20 | 0.040 | 12-25 | **0.070** | **20** | NP | NP | 0.030 | 60 | NP | NP | NP | NP |
| **NS_MID** | **0.180** | **10** | 0.050 | 10-25 | 0.065 | 25 | NP | NP | NP | NP | 0.050 | 25-30 | 0.075 | 15 |
| **NS_LOW** | 0.050 | 15 | NP | NP | 0.040 | 15 | **0.060** | **25** | 0.050 | 30-45 | NP | NP | NP | NP |
| **ARE** | 0.050 | 10 | NP | NP | 0.038 | 10 | 0.025 | 30-40 | NP | NP | 0.025 | 15-35 | **0.060** | **35** |
| **ARE_HIGH** | 0.029 | 6 | 0.037 | 80 | 0.027 | 20-32 | NP | NP | 0.027 | 65 | **0.075** | **15** | 0.025 | 35 |
| **ARE_MID** | **0.050** | **10** | 0.030 | 40 | 0.040 | 12 | 0.037 | 15-30 | 0.040 | 30 | 0.040 | 12 | 0.037 | 20-25 |
| **ARE_LO W** | 0.038 | 9 | 0.027 | 10 | 0.090 | 5 | 0.036 | 27-40 | NP | NP | 0.036 | 25-30 | **0.063** | **27** |
| **JSI** | **0.150** | **5** | NP | NP | 0.075 | 20 | 0.037 | 12 | 0.030 | 12 | NP | NP | NP | NP |
| **JSI_HIGH** | 0.013 | 10 | NP | NP | **0.290** | **18** | 0.050 | 10 | 0.015 | 18 | 0.075 | 15 | 0.100 | 15 |
| **JSI_MID** | 0.100 | 5 | NP | NP | **0.310** | **5** | NP | NP | NP | NP | 0.050 | 10 | NP | NP |
| **JSI_LOW** | **0.430** | **5** | NP | NP | 0.075 | 20 | NP | NP | NP | NP | NP | NP | NP | NP |
| **CAL** | 0.049 | 6 | NP | NP | **0.053** | **6** | 0.030 | 12 | NP | NP | 0.025 | 20 | 0.025 | 15 |
| **CAL_HIG H** | **0.120** | **3** | NP | NP | 0.040 | 10 | 0.090 | 15 | 0.024 | 35 | NP | NP | 0.060 | 17 |
| **CAL_MID** | 0.028 | 30 | NP | NP | 0.075 | 10 | 0.025 | 30 | 0.050 | 110 | **0.090** | **25** | 0.025 | 25 |
| **CAL_LO W** | **0.120** | **3** | 0.05 | 100-125 | 0.090 | 5 | 0.050 | 5 | - | - | NP | NP | 0.060 | 10 |

<sup>1</sup>DP = Density peak/ <sup>2</sup>PD = Peak distance/ <sup>3</sup>NP = no peak, flat curve.

Between 40% and 81% of drainage components were classified as inadequate or partially adequate, and all study areas presented a quantitative deficit of drainage infrastructure (Fig 3). Density analyses showed no clear pattern for distance from individuals with anti-*Leptospira* antibodies and problematic drainage components among study areas and sub-areas. Nova Sussuarana and Arenoso have storm drainage systems in worse condition uphill compared to downhill (Fig 3a,b), which is the opposite observed for inadequate streets. Problematic sewerage components are evenly distributed among sub-areas, although coverage of sewerage increases from uphill to downhill (Fig 3). Individuals with anti-*Leptospira* antibodies tended to live within 25 m of problematic sewerage components (Fig 4). Generally, problematic sewerage components appear to be the most relevant risk factor in terms of density of individuals with anti-*Leptospira* antibodies after inadequate streets (Table 3). Sub-area analyses showed heterogeneous patterns among study areas.

Polluted streams pass through all areas and sub-areas, with lengths ranging from 87 to 203 m (Fig 3). Individuals with anti-*Leptospira* antibodies generally reside within 50 m of a polluted stream. There were no prominent density peaks, but higher densities occurred between 12 and 25 m (Fig 4), but lower than the observed for inadequate streets and problematic sewerage components. The number of open sewage points located outside the stream vary from three to nine across the study areas (Fig 3). Density curves are relatively flat, indicating a weaker spatial concentration of individuals with anti-*Leptospira* antibodies than that observed for inadequate streets and problematic sewerage components (Fig 4). At sub-area level, higher densities were observed in uphill or intermediate sub-areas, as the density curves in the lower sub-areas are predominantly flat and close to zero (Fig 5).

Across the study areas, there are between 16 to 22 open waste dumping sites (Fig 3), with average volumes ranging from 4.3 m³ and 9.8 m³, and total volumes between 68 m³ and 162 m³ across the study areas. Density curves show relatively lower peaks compared to street or sewerage-related variables (Fig 4). Across sub-areas, the lower densities of individuals with anti-*Leptospira* antibodies coincided with sub-areas where there were less waste dumping sites (Fig 5).

There are between nine and fifteen vacant lots across the study areas (Fig 3). Within each study area, between 19 and 41% of waste sites coincided with vacant lots, indicating a spatial overlap between these variables. Density curves show higher densities between 15 m and 35 m in three out of four areas, indicating a higher probability of individuals with anti-*Leptospira* antibodies within this distance range from vacant lots (Table 3). At the sub-area level, the highest densities were observed in the upper sub-area of Jardim Santo Inacio, where most waste points cluster on vacant lots (Fig 5c).

### 3.2. Relative elevation of households

The density analysis regarding relative elevation of households shows that in all study areas, the individuals with anti-*Leptospira* antibodies tend to live at the lowest elevations, with peaks at around 2 to 3 m (Fig 6).

**Fig 6.**
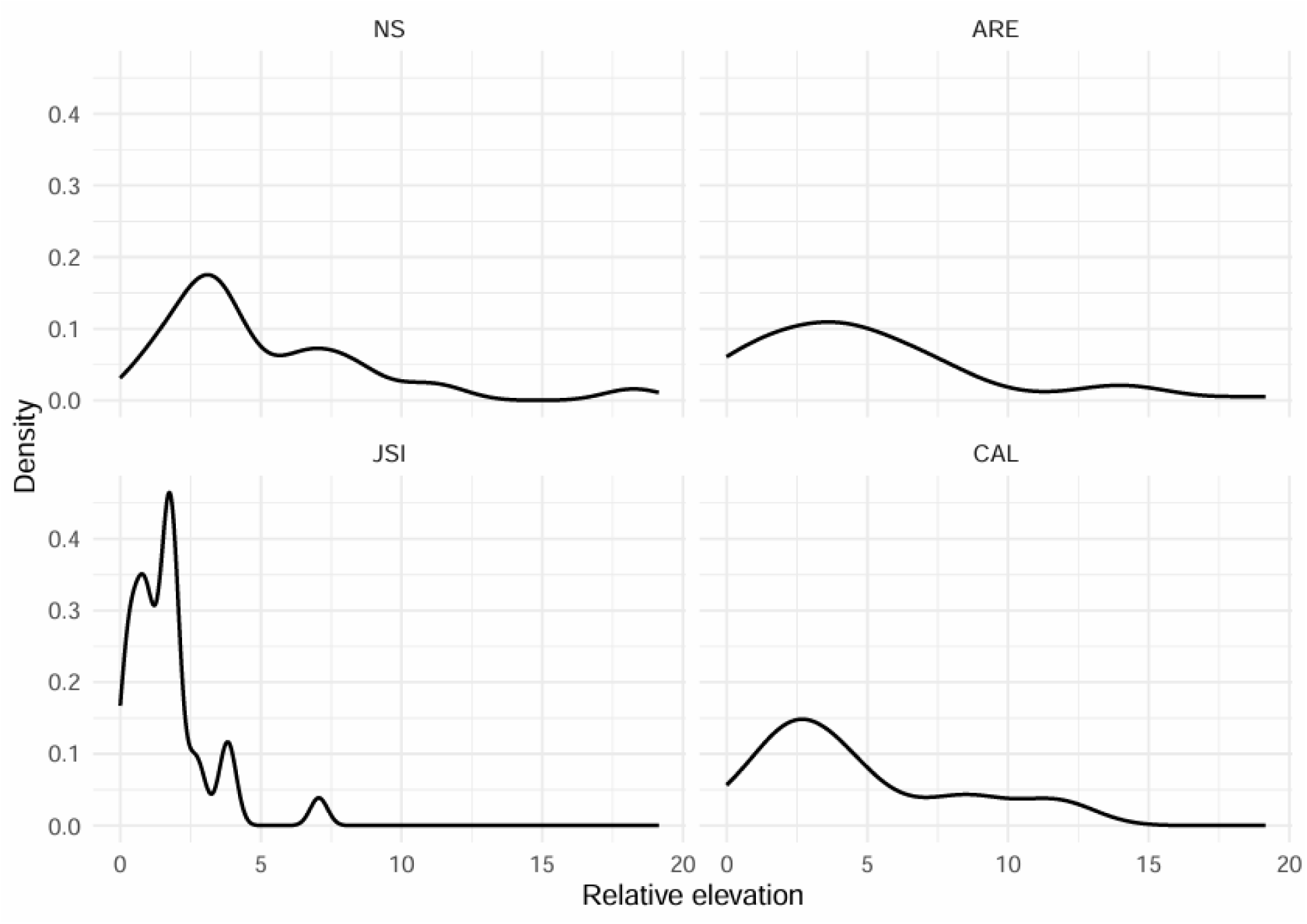
Probability density of relative household elevation among seropositive individuals in the four study areas.

## 4. DISCUSSION

### 4.1. Sewerage infrastructure as an overlooked exposure pathway

This study developed and implemented a novel methodology to characterise sanitation, drainage and waste infrastructure and related problems in urban deprived areas. The approach enabled the identification of problematic sewerage infrastructure as one of the risk factors most consistently close to individuals with anti-*Leptospira* antibodies, highlighting exposure pathways beyond streams and open sewage points. Across the four study areas, infected individuals tended to reside closer to inadequate and partially adequate sewerage components than to streams or open sewage points, suggesting that sewerage infrastructure itself may play an important role in environmental exposure.

Density analysis highlights the importance of sewage-related risk factors. Individuals with anti-*Leptospira* antibodies tended to live near risk factors. Across the four study areas, this distance is approximately 10 m for inadequate or partially adequate sewerage. The density peak for sewerage risk factors is higher than for streams and open sewage points. Previous studies have consistently reported increased risk among residents living closer to sewage-contaminated environments, particularly in relation to ‘open sewers’ [10,34,35]. However, the open sewers in previous studies in Salvador (Brazil) do not have the same characteristics as the streams in this study as they were more accessible to pedestrians and shallower than those found in the study areas here.

The relatively small number of open sewage points, compared to sewerage components, likely results in longer average distances, leading to a more spatially dispersed pattern. Individuals may reside up to 100 m from an open sewage point, at least double the maximum distance observed for sewerage components. Two key points emerge from this: 1) The higher frequency of problematic sewerage components compared to open sewage points may increase exposure to sewage via sewerage infrastructure; 2) The small number of open sewage points may lead to an underestimation of exposure, necessitating careful interpretation of density analysis, particularly when sample sizes are small.

It is important to note even a single open sewage point represents a potential exposure site where individuals may come into contact with contaminated water, either directly at the source or indirectly through contamination that spreads from that point. Therefore, the presence of even few dispersed sewage points can still pose significant health risks, highlighting the importance of not underestimating exposure based solely on their number or density. Furthermore, open sewage points may be acting as a more diffuse risk factor that promote the spread of *leptospires* in the environment.

The density curves for streams follow a similar pattern to those of open sewage points. However, the underlying reasons for this likely differ. Streams traverse the study areas, creating more points for distance calculations. The spatial distribution of households relative to streams appears consistent across the four study areas, whereas sewerage components exhibit greater variability. Additionally, households in the central part of the study areas tend to be closer to streams, but elevation varies in two directions: (1) longitudinally, from upstream to downstream, and (2) transversally, from higher areas (farther from the stream) to lower areas (closer to the stream). This suggests that factors beyond proximity to streams influence leptospirosis transmission and should be analysed in combination with other variables, such as elevation [31].

The findings from this study indicate that inadequate or partially adequate sewerage may be more relevant for leptospirosis transmission than sewage-contaminated streams and open sewage points in these communities. One possible explanation is the greater frequency and wider spatial distribution of sewerage components. Additionally, sewage spillage tends to create stagnant pools in unpredictable locations, making them less noticeable and harder to avoid. In contrast, streams and open sewage points are more visibly hazardous, meaning people are more likely to perceive and avoid them.

### 4.2. Elevation, inadequate streets and storm drainage

The density analysis shows that individuals with anti-*Leptospira* antibodies tend to reside at lower relative elevations in all four study areas. This finding is consistent with previous studies that have identified the individuals who live at lower elevations are at higher risk for Leptospiral infections [17,31]. This supports the hypothesis that hydrological processes play a key role in transmission [31]. Previous studies had also found that rat abundance had a stronger effect at higher elevations, where water runoff is lower, suggesting that in lower areas, water dispersal plays a more dominant role in transmission. In contrast, at higher elevations, where hydrological factors are less relevant, contamination may be more localised and therefore more influenced by rat activity [31].

The analyses according to sub-areas reveal that inadequate streets, a variable proposed as a potential risk factor in this study, are more common in the lower sub-areas of three out of four study areas. This is possibly caused by the poor drainage system since upstream combined with the higher water runoff in lower areas. Among individuals with anti-*Leptospira* antibodies, density peaks at around 10 metres from inadequate streets. If these individuals are more likely to reside at lower elevations and in proximity to inadequate streets, further investigation is necessary to determine whether household elevation and street conditions act as confounding factors or independent risk factors.

Streets may be a significant source of contamination. Previous studies have detected *Leptospira* in streets affected by sewage contamination [13]. Theoretically, streams and open sewers would have a higher pollution load, particularly in terms of sewage discharge, and potentially a higher *Leptospira* concentration compared to streets. However, inadequate streets could present a greater risk in terms of frequency of contact, and it could be more relevant in upper areas where contamination dispersed from streams is less relevant and contamination in the streets would be more important.

Residents might not come into direct contact with streams, particularly those living farther away or at higher elevations relative to the stream. For these individuals, the stream may not be the most relevant risk factor in terms of direct exposure, but streets would be. This supports the hypothesis raised that hydrological profiles play an important role [31]. If rat abundance is more relevant in upper areas, it is possible that transmission primarily occurs through contaminated streets, particularly after rainfall events.

The drainage system plays an important role in water runoff. In addition to the high proportion of problematic drainage components identified in this study, particularly in the upper sub-areas, all study areas exhibited a quantitative deficit of drainage infrastructure. Drainage system failures upstream may slow down water flow towards lower areas, leading to localised flooding and increasing the likelihood of pedestrian contact with contaminated water during or after rainfall events. This could help explain leptospirosis transmission in upper areas, not necessarily flood-prone zones, but areas affected by drainage infrastructure failures. Previous studies have similarly highlighted the role of outdated storm drainage systems in increasing human exposure to contaminated water and the importance of maintenance to prevent flash floods [22,36].

Therefore, these findings suggest that elevation, street conditions and drainage infrastructure interact and influence environmental exposure pathways. Further investigations should integrate hydrological characteristics when designing studies and selecting study areas to better understand how water movement influences leptospirosis transmission.

### 4.3. Open solid waste dumping sites and vacant lots

Individuals with anti-*Leptospira* antibodies are mostly located within 50 m of waste dumps or vacant lots but no clear pattern of prominent peaks emerged. Furthermore, 50 m appears to be an important cut-off point in three out four areas, indicating a very low probability of individuals with anti-*Leptospira* antibodies residing beyond this distance from points of waste accumulation and vacant lots. Previous studies have identified accumulated waste as an important environmental factor associated with leptospirosis risk and rat abundance [31,37]. Waste accumulation may provide food resources and shelter for rodents, thereby indirectly increasing opportunities for environmental contamination. The diffuse spatial patterns observed in this study suggest that waste accumulation may influence transmission through broader ecological processes rather than acting as highly localised exposure points.

Vacant lots may play a complementary role. A substantial proportion of waste accumulation points coincided with vacant lots, providing favourable conditions for rat proliferation. The literature does not explicitly consider vacant lots as a risk factor for leptospirosis as frequently as open waste dumping sites [10,35,38]. Residence bordering vacant lot has been associated with small mammal assemblages, including leptospirosis-infected rats [39].

### 4.4. Study Limitations and Contributions

This study has some limitations. Density analyses identify spatial patterns but do not establish causal relationships. Some variables, particularly open sewage points, presented relatively few observations, which may influence the interpretation of density patterns. Infrastructure condition was assessed through external visual inspection, meaning that internal obstructions or functioning may not have been detected and quantified. It may involve residual subjectivity despite the use of standardised protocols. While subjectivity cannot be fully eliminated in this type of assessment, further work is needed to explore how it can be reduced. In addition, vacant lots were identified but not characterised by specific environmental attributes that may influence rats’ activity.

This was a cross-sectional study, and we could not therefore assess how the importance of risk factors may change over time. Future studies should prioritise collecting longitudinal data which would permit integration of antecedent rainfall as a factor influencing the importance and role of different risk factors. In addition, because the study areas were purposively selected, the findings may be applicable to similar urban settings but not generalised to all urban informal settlements. However, the methodology may be transferable to a broader range of low-income urban settings.

Despite these limitations, this study provides novel approach and a high-resolution assessment of sanitation, storm drainage, and waste-related infrastructure in urban informal settlements. Key strengths of this study are the comprehensive assessment framework and the use of short-distance measurements with high precision, leading to more reliable spatial analyses. By evaluating environmental risk factors, it highlights infrastructure characteristics and potential exposure pathways that needs further investigation.

This study has shown that a detailed assessment of infrastructure risk factors for exposure *Leptospira* can improve the understanding of potential *Leptospira* exposure pathways in urban informal settlements therefore where more action is required. Overall, inadequate streets and problematic sewerage components showed the highest density peaks and shortest distance to individuals with anti-*Leptospira* antibodies. In contrast, streams, open sewage points, waste dumping sites and vacant lots presented more diffuse spatial patterns. This indicates that environmental and infrastructure risk factors work at different scale and may warrant different approaches to reduce threats. The findings suggest that infrastructure deficiencies differ in their potential contribution to environmental exposure and should not be considered equally important when prioritising interventions. High-precision georeferencing helps to identify priority areas for improvements and guide future epidemiological investigations.

## CRediT authorship contribution statement

**Ana Maria Nascimento Silva**: Conceptualization, Data curation, Formal analysis, Investigation, Methodology, Visualization, Writing – original draft, Writing – review & editing. **Juliet Oliveira Santana**: Conceptualization, Data curation, Investigation, Methodology, Visualization, Writing – review & editing. **George Gonçalves Machado**: Conceptualization, Data curation, Investigation, Methodology, Writing – review & editing. **Fábio Neves Souza**: Conceptualization, Methodology, Resources, Writing – review & editing. **Daiana Santos de Oliveira**: Data curation, Investigation, Writing – review & editing. **Fabiana Almerinda G. Palma**: Conceptualization, Resources, Writing – review & editing. **Priscilla Elizabeth Ferreira dos Santos**: Resources, Investigation. **Paulo Ricardo Dias Pimentel**: Data curation, Investigation. **Cleber Cremonese**: Conceptualization, Project administration, Resources, Supervision. **Federico Costa**: Conceptualization, Funding acquisition, Methodology, Project administration, Supervision, Writing – review & editing. **Rodolfo B. Nobrega**: Conceptualization, Methodology, Supervision, Writing – review & editing. **Guy Howard**: Conceptualization, Funding acquisition, Methodology, Project administration, Resources, Supervision, Writing – review & editing.

## Acknowledgements

We would like to thank the residents in the study areas, participants and community leaders.

## Declaration of Generative AI use

During the preparation of this work, the authors used ChatGPT (OpenAI) to assist with language editing, improving clarity, and enhancing the readability of the text. After using this tool, the authors reviewed, edited, and verified all content and take full responsibility for the content of the published article.

## Data Availability Statement

The data is managed in accordance with the ethical standards, detailed in the methodology section. Due to the inclusion of personal information from survey participants, the datasets used and analysed during the current study cannot be publicly shared. Interested researchers can contact Dr Federico Costa to obtain the raw data, clearly stating the purpose of their study including the protocol and analysis plan.

## Supporting Information

**S1 Table**. Criteria for segment definition.

**S1 Figure**. Example of the subdivision of Study Area 1 into hydrological sub-areas following the flow direction: (a) upper, (b) intermediate, and (c) lower. Blue lines indicate the sub-catchment boundaries used to delineate the sub-areas.

**S1 Checklist.** STROBE checklist for the cross-sectional analysis.

